# The Longevity Landscape: Value Creation for Healthy Aging

**DOI:** 10.1101/2024.05.28.24308017

**Authors:** Wasu Mekniran, Odile-Florence Giger, Elgar Fleisch, Tobias Kowatsch, Mia Jovanova

## Abstract

**Background:** The global aging population underscores a critical need to tackle accompanying health and economic challenges, at all levels of society. This *All-of-Society* approach emphasizes the involvement of various stakeholders—governments, NGOs, researcher centers, private companies, local communities, and opinion leaders—to collectively promote healthy aging. However, how stakeholders enable healthy longevity remains unclear.

**Objective:** This study examines how global stakeholders (governments, NGOs, researcher centers, private companies, local communities, and opinion leaders) create value towards healthy longevity. We identify the healthy longevity dimension of stakeholders’ value propositions and examine alignment between their propositions as an indicator of shared goals.

**Methods:** Following the *All-of-Society* approach, we analyzed the healthy longevity aspects of value propositions among the six classes of stakeholders (*N*=128). We (1) employed semantic topic modeling to identify the primary value proposition topics as related to healthy longevity and (2) computed proposition alignment using similarity networks.

**Results:** Our analysis revealed varying degrees of alignment between stakeholders’ healthy longevity propositions, with the lowest alignment observed for local communities and researcher centers.

**Conclusions:** Findings underscore a key need to strengthen synergies between academic and community-based initiatives to promote translational science and highlight opportunities for strategic partnerships in the evolving healthy longevity field.

**What is already known on this topic:** The National Academy of Medicine’s *All-of-Society* approach advocates for multi-stakeholder engagement towards healthy longevity, but specific stakeholder contributions, and their alignment toward shared goals, are poorly understood.

**What this study adds:** To our knowledge, this study is the first to provide empirical evidence into the value propositions of healthy longevity stakeholders on a societal scale. It highlights key areas where multi-stakeholder collaboration can be strengthened—particularly between academic and local community initiatives—and proposes five strategies to strengthen collaboration.

**How this study might affect research, practice, or policy:** Prioritizing (1) community-based participatory research, (2) translating healthy aging-related research findings into accessible resources, (3) prioritizing equity in intervention delivery, (4) establishing community advisory boards, and (5) developing knowledge translation and training programs, could potentially better align academic and community efforts towards more aligned, equitable and effective healthy longevity initiatives.

## INTRODUCTION

The global demographic change toward an aging population, combined with the surge of non-communicable diseases (NCDs), presents key challenges in our healthcare systems (WHO, 2020). This challenge is two-fold: on an individual scale, older adults increasingly contend with NCDs as they age, while at a societal level, healthcare systems face escalating costs. Notably, 74% (41 out of 55.4 million) global deaths in 2019 were attributed to NCDs (WHO, 2023). In parallel, global healthcare expenditure is projected to surge from $9.1 trillion in 2020 to $11 trillion by 2026 (IHME, 2023). These alarming numbers underscore the need to prioritize healthy aging.

Addressing these complex challenges calls for healthy longevity initiatives, ensuring individuals maintain their functional health as they age (Bautmans *et al*., 2022). Recognizing this urgency, WHO (2020) designated 2021–2030 as the Decade of Healthy Aging, and the National Academy of Medicine (NAM) proposed the *All-of-Society* approach, with different sectors coming together to tackle healthy aging, at a societal level (WHO, 2020; National Academy of Medicine, 2022). Yet, implementing a multi-stakeholder approach is complex; and no prior work has evaluated stakeholder roles. This study aims to address this gap and identify opportunities to build stakeholder synergies.

Drawing from public health and business research, a helpful method to understand a stakeholder’s role is by analyzing their value proposition (Payne and Frow, 2014; Gassmann, Frankenberger and Choudury, 2020). A value proposition captures what a stakeholder offers and to whom (Steinhöfel, Kohl and Orth, 2016) and serves as a foundation for multi-stakeholder collaboration. Theoretically, stronger value alignment between stakeholders suggests shared objectives and fosters partnerships to address complex challenges like global aging, ensuring that all stakeholders are working towards the same objectives. This shared focus from a complementary value proposition could enhance cooperation, and support a unified approach to tackling issues, thereby increasing the likelihood of sustainability of outcomes (Cozzolino and Geiger, 2024). However, no work to date has examined the value propositions of stakeholders in the longevity landscape, which is crucial to understanding their individual roles and collaborative dynamics (Lingens, Seeholzer and Gassmann, 2022). Thus, this study aims to (a) identify the healthy longevity value propositions of key stakeholders and (b) analyze their alignment as an index of potential collaboration. We examine the following questions:

RQ1: What are the value propositions of healthy longevity stakeholders?

RQ2: How aligned are the value propositions of healthy longevity stakeholders?

By addressing these questions, we aim to identify potential gaps in value alignment and discuss recommendations towards more collaborative healthy longevity efforts.

## METHODS

To identify and analyze the value propositions of key classes of stakeholders (governments, NGOs, researcher centers, private companies, local communities, and opinion leaders) in healthy longevity, we conducted a content analysis and applied computational social science tools. We identified academic and non-academic stakeholders (*N*=128) using pre-defined search terms and extracted their value propositions using a combination of databases (i.e., Web of Science, PitchBook, and Crunchbase). For details on the search strategy, data inclusion, methods details, and analysis plan, refer to Appendix S1. Next, we applied semantic topic modeling, a data-driven text analyses technique (Valdez, Pickett and Goodson, 2018) to identify the primary themes in the healthy longevity value propositions (RQ1) and network analysis (Wasserman and Faust, 1994) to assess alignment between stakeholders’ value propositions (RQ2). Building on these analyses, we propose recommendations to strengthen stakeholders’ value alignment. No ethics approval was required for this study, as it involves the analysis of publicly available data and no human subjects or personally identifiable information.

## RESULTS

### Value creation among stakeholders in the healthy longevity landscape

Using semantic topic modeling, we categorized the main themes of the healthy longevity aspects of the value propositions of the six classes of stakeholders (RQ1):

In our sample, governmental organizations focus on developing policies that create supportive environments for older adults and establish evaluation guidelines for standardized program evaluation. NGOs focus on allocating longevity-focused resources among stakeholders and developing healthy aging programs. Research centers investigate aging biology and develop interventions for NCD prevention and management. Private companies in the longevity sector focus on personalized, digital health analytics to prevent and treat age-related diseases. Local community supports equitable aging through targeted education, family support initiatives, and advocates for aging-specific healthcare services for diverse populations. Opinion leaders promote longevity innovations through outreach activities, including disseminating research on age reversal methods. See Figure S2 for main themes per stakeholder.

### Value alignment among stakeholders in the longevity landscape

To examine how likely are stakeholders to share common healthy longevity related values, we analyzed the similarity of their value propositions. We created an undirected network where stakeholders are represented as nodes, and the connections, or edges, between them are based on how similar their value propositions are to each other (Figure 1, A). Similarity was measured using a widely used pairwise-cosine similarity score (Valdez, Pickett and Goodson, 2018). By summing up these similarity values for each stakeholder across the network, we calculated their “node strength,” which indicates how closely their values align with those of other stakeholders. Higher node strength indicates stronger alignment with the overall values of the network. For more detailed information on the measurement process, refer to Appendix S1.

**Figure 1.**
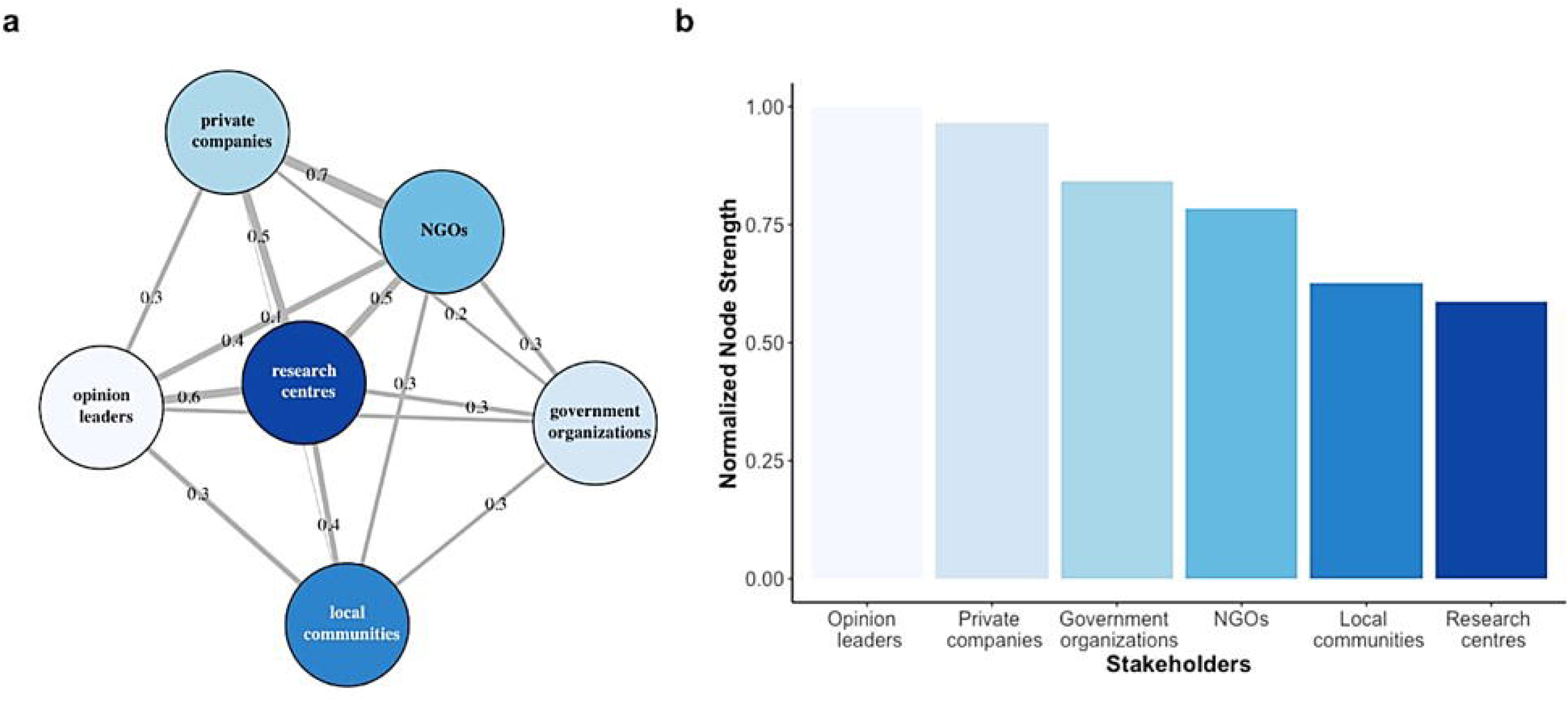
Healthy longevity value proposition network, where nodes are stakeholders and edges present pairwise cosine similarity scores (A). Node strength distribution for stakeholders in healthy longevity (B). *Note*: node strength is normalized (0-1).

We observed wide variability in stakeholders’ healthy longevity value proposition alignment (normalized node strength range: 0.59-1) (Figure 2, B). Opinion leaders, private companies, NGOs, and government organizational, showed high to medium alignment with all other stakeholders, with shared overarching goals to fund, develop, implement, evaluate, and disseminate healthy aging programs, while local communities and research centers displayed lower levels of alignment (relative to all other stakeholders), with normalized node strengths of 0.63 and 0.59, respectively, implying lower alignment with other stakeholders’ goals (Figure 2, B). Researcher centers focus on innovative aging-disease related research, while local communities focus on equitable healthy aging advocacy and reducing disparities in healthy aging.

## DISCUSSION

This report analyzes how global stakeholders create value towards healthy longevity with a systematic approach. We identified the healthy longevity-related value propositions of 128 stakeholders—government organizations, NGOs, research centers, private companies, local communities, and opinion leaders, through semantic topic modeling and network analyses and examined their alignment. Our findings uncovered both consistent and inconsistent values. Specifically, we observed stronger value alignment among opinion leaders, private companies, NGOs and government organizations, in funding, developing, implementing, evaluating, and disseminate healthy aging programs, but weaker alignment for local communities and research centers relative to all other stakeholders.

Importantly, our findings suggest that while research centers may drive biological and behavioral innovations and advancements to promote healthy aging, conflicts may arise over accessibility, affordability, and equity in delivering these innovations to individuals and communities who need them most (Fernandez-Moure, 2016). Thus, strengthening the alignment between local and academic stakeholders is a first step to foster more coordinated initiatives that do not exacerbate existing health inequalities; and are better suited to the needs of local communities. To this end, we propose five strategies for value alignment—particularly to strengthen accessibility, affordability, and equity in developing and delivering scientific advancements for healthy longevity:

1. **Community-Based Participatory Research**: Actively involving community members in longevity research development processes.
2. **Translating Healthy Aging-related Research Findings into Accessible Resources**: Ensuring research findings are accessible and tailored to community needs and address specific concerns or interests within the community.
3. **Prioritizing Equity in Healthy Aging Intervention Delivery:** Tailoring interventions to address specific barriers: i.e., socioeconomic, or cultural factors that may affect intervention access.
4. **Establishing Community Advisory Boards:** Creating longevity council boards where residents provide input on healthy aging research initiatives.
5. **Developing Knowledge Translation and Training Programs**: Empowering community members with information related to healthy aging, i.e., online training modules or workshops designed to promote healthy aging behaviors.

These proposed strategies reshape current stakeholder priorities with a focus on equity in healthy aging and aim to ensure that scientific progress from research centers reaches diverse populations outside the lab. Together, while our analysis of healthy longevity value propositions contributes to new opportunities for stakeholder partnerships, it is also important to acknowledge that our sampling may not capture all emerging stakeholders in the longevity landscape and does not focus on payers and payees. The novelty of the healthy longevity field means that scientific publications and financial data are limited and may omit additional relevant sources. Future work could address these limitations by employing more systematic sampling approaches at scale and conducting expert interviews.

Overall, this work improves our understanding of stakeholder values and collaboration dynamics in the healthy longevity landscape and underscores and opportunity to create synergies between local communities and research centers to address healthy aging. We emphasize the importance of community-based participatory research, accessible delivery of aging interventions, establishment of healthy aging community advisory boards, and implementation of knowledge translation programs as key steps towards healthy longevity equity.

## Data Availability

All data produced in the present study are available upon reasonable request to the authors

## Contributions

WM, TK and MJ contributed to the conceptualization of this research. WM and OFG conducted the qualitative data collection and content analysis. MJ conducted the quantitative analyses. WM wrote the research protocol and the first version of the manuscript. MJ, EF, and TK provided feedback on the manuscript. All authors reviewed and edited the manuscript and approved the final version.

## Patients and Public Involvement

This research did not involve patients or the public directly.

---

**Figure S2.**
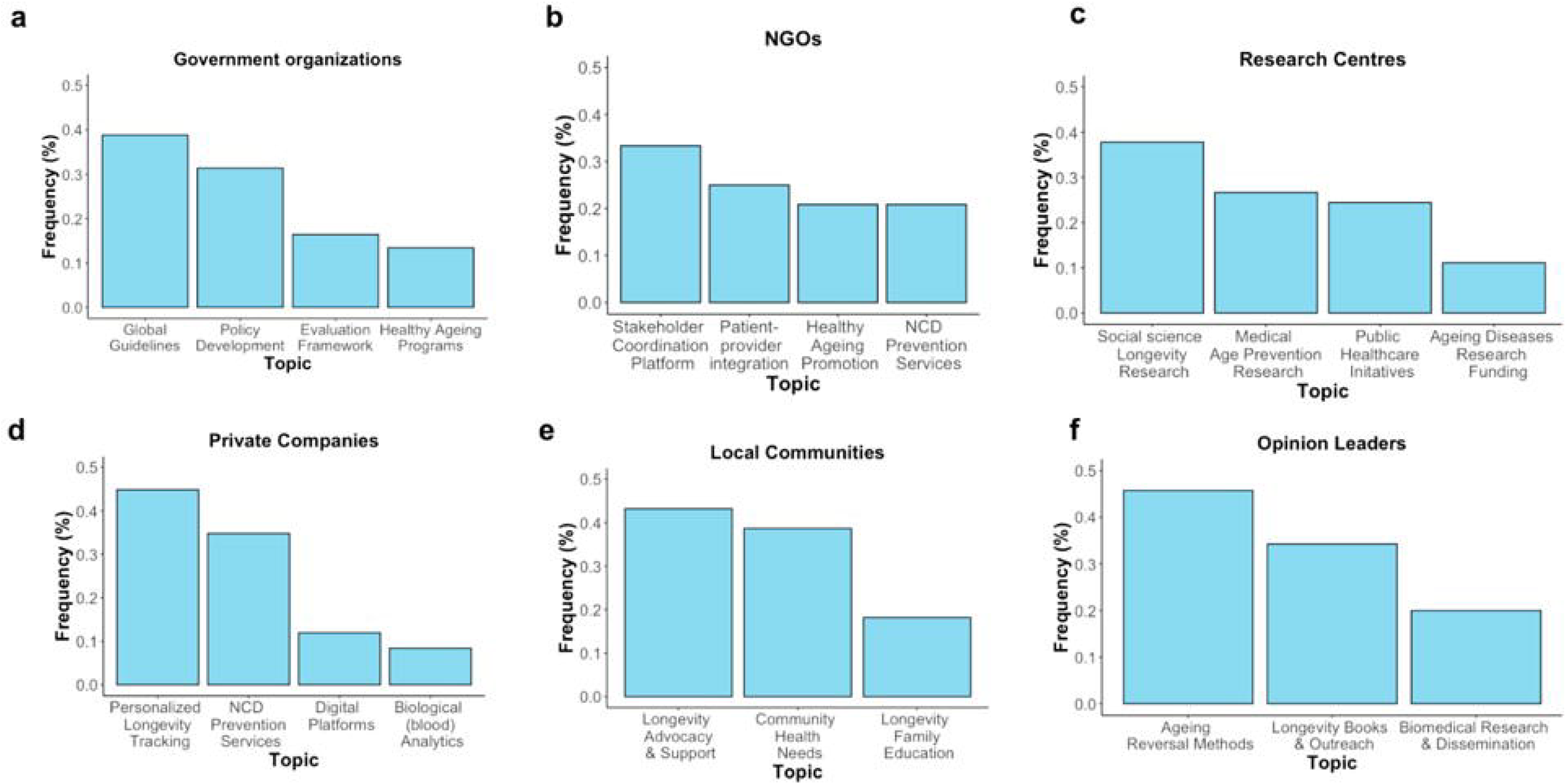
Key topics identified in stakeholders’ healthy longevity related value proposition statements *(N=128)* using semantic topic modeling.

## REFERENCES

Bautmans, I. et al. (2022) ‘WHO working definition of vitality capacity for healthy longevity monitoring’, The Lancet Healthy Longevity, 3(11), pp. e789–e796. Available at: 10.1016/S2666-7568(22)00200-8.

Cozzolino, A. and Geiger, S. (2024) ‘Ecosystem disruption and regulatory positioning: Entry strategies of digital health startup orchestrators and complementors’, Research Policy, 53(2), p. 104913. Available at: 10.1016/j.respol.2023.104913.

Crunchbase (2024) ‘Crunchbase Inc.’ Available at: https://www.crunchbase.com/home (Accessed: 7 May 2024).

Ebadi, A. and Schiffauerova, A. (2016) ‘How to boost scientific production? A statistical analysis of research funding and other influencing factors’, Scientometrics, 106(3), pp. 1093–1116. Available at: 10.1007/s11192-015-1825-x.

Fernandez-Moure, J.S. (2016) ‘Lost in Translation: The Gap in Scientific Advancements and Clinical Application’, Frontiers in Bioengineering and Biotechnology, 4. Available at: 10.3389/fbioe.2016.00043.

Fried, L.P., Wong, J.E.-L. and Dzau, V. (2022) ‘A global roadmap to seize the opportunities of healthy longevity’, Nature Aging, pp. 1–4. Available at: 10.1038/s43587-022-00332-7.

Gassmann, O., Frankenberger, K. and Choudury, M. (2020) The Business Model Navigator: 55+ models that will revolutionise your business. Second Edition. New York: Pearson.

IHME (2023) ‘Financing Global Health’. University of Washington. Available at: https://vizhub.healthdata.org/fgh/.

Jelodar, H. et al. (2019) ‘Latent Dirichlet allocation (LDA) and topic modeling: models, applications, a survey’, Multimedia Tools and Applications, 78(11), pp. 15169–15211. Available at: 10.1007/s11042-018-6894-4.

Lingens, B., Seeholzer, V. and Gassmann, O. (2022) ‘The architecture of innovation: how firms configure different types of complementarities in emerging ecosystems’, Industry and Innovation, 29(9), pp. 1108–1139. Available at: 10.1080/13662716.2022.2123307.

National Academy of Medicine (2022) Global Roadmap for Healthy Longevity. Washington, D.C.: National Academies Press, p. 26144. Available at: 10.17226/26144.

Payne, A. and Frow, P. (2014) ‘Developing superior value propositions: a strategic marketing imperative’, Journal of Service Management. Edited by P. Bo Edvardsson and Professor Philipp Klaus, 25(2), pp. 213–227. Available at: 10.1108/JOSM-01-2014-0036.

PitchBook (2024) ‘Venture Capital, Private Equity and M&A Database’. Available at: https://pitchbook.com/ (Accessed: 7 May 2024).

Silge, J. and Robinson, D. (2017) Text mining with R: a tidy approach. First edition. Beijing Boston Farnham Sebastopol Tokyo: O’Reilly.

Steinhöfel, E., Kohl, H. and Orth, R. (2016) Business Model Innovation: A Comparative Analysis.

Valdez, D., Pickett, A.C. and Goodson, P. (2018) ‘Topic Modeling: Latent Semantic Analysis for the Social Sciences’, Social Science Quarterly, 99(5), pp. 1665–1679. Available at: 10.1111/ssqu.12528.

Wasserman, S. and Faust, K. (1994) Social Network Analysis: Methods and Applications. Cambridge: Cambridge University Press (Structural Analysis in the Social Sciences). Available at: 10.1017/CBO9780511815478.

WHO (2020) Decade of healthy ageing: baseline report. Geneva: World Health Organization. Available at: https://apps.who.int/iris/handle/10665/338677 (Accessed: 12 September 2022).

WHO (2023) Non communicable diseases Key Facts. Available at: https://www.who.int/news-room/fact-sheets/detail/noncommunicable-diseases (Accessed: 12 March 2024).

