## Supplement for "The Longevity Landscape: Value Creation for Healthy Aging"

### APPENDIX

*Search Strategy and Data Collection*

Following the *All-of-Society* approach, we aimed to identify key stakeholders —government organizations , NGOs, researcher centers, private companies, local communities, and opinion leaders—most involved in the healthy longevity landscape (Fried, Wong and Dzau, 2022). Our search strategy involved utilizing multiple databases. We utilized Web of Science to capture stakeholders that devote the top research funding towards longevity science, including top-funded longevity-focused research centers and sponsoring government organizations and NGOs. Additionally, PitchBook and Crunchbase were used to ensure comprehensive coverage of non-academic stakeholders, such as private companies, local communities, and opinion leaders, that generate the top funding and activity in the healthy longevity sphere (Crunchbase, 2024; PitchBook, 2024). These databases specifically allow for a backward search approach based on the total funding devoted to industry (i.e., longevity programs) and for specific individuals, i.e., based on their participation in industry-related events (i.e., global longevity congresses). To be included in the analysis, stakeholders needed to meet the following predefined inclusion criteria: (a) involvement in initiatives related to healthy longevity and (b) articulation of value propositions or objectives explicitly aligned with promoting healthy aging or longevity (WHO, 2020).

To identify key stakeholders, we extracted entries based on our search strategy shown in Table S1, and we selected the top 20 results in each category based on funding, as these are likely to be the top-funded stakeholders who drive the development trajectory of the healthy longevity landscape (Ebadi and Schiffauerova, 2016). During the screening of key stakeholders, we further identified relevant stakeholders using a snowball sampling approach and included them to be screened for eligibility. Next, two coders screened all entries for eligibility. They extracted the value propositions of each eligible stakeholder as presented on their respective scientific, financial data, or personal websites, with a third coder resolving any discrepancies. Our final sample included *N=128* stakeholders in total (governmental organizations = 8; multilateral organizations and NGOs = 9; research centers = 11; private companies = 79; communities = 11; opinion leaders = 10). See Table S1 for the search strategy and example stakeholders included in our sample and Figure S1 for the stakeholder eligibility flowchart.

**Table S1. Search strategy.**

| Stakeholder | Database | Criteria | Search Terms | Samples |
|---|---|---|---|---|
| (1) Government organizations<br>(2) NGOs<br>(3) Researcher centers | Web of Science | Total funding devoted to healthy longevity scientific research | Topic: ("Healthy Ageing" OR "Healthy Longevity") | (1) UN, WHO, EU<br>(2) Wellcome Trust, AGE-WELL<br>(3) MIT AgeLab, Buck Institute for Research on Aging |
| (4) Private companies | Crunchbase/ PitchBook | Total funding amount devoted to longevity-programs, communities, or private enterprises | Keywords: Healthy Longevity OR Healthcare OR Biological age;<br><br>Industry function: Health care | (4) Novartis, MassMutual, Noom, TruDiagnostic, Insilico Medicine, Apollo Health Ventures |
| (5) Communities | | | Keywords: Healthy Longevity OR Healthcare OR Biological age;<br><br>Industry function: Communities | (5) Cedars Sinai, MaineHealth, Heartland Family Service |
| (6) Opinion leaders | | Individual participation in the highest number of global healthy longevity events | Event function: Longevity Leaders World Congress OR International Longevity Policy and Governance Summit | (6) David Sinclair, Aubrey de Grey, Jim Mellon, James Peyer |

**Figure S1.** Flow diagram and exclusion criteria for identifying key stakeholders in the healthy longevity landscape.

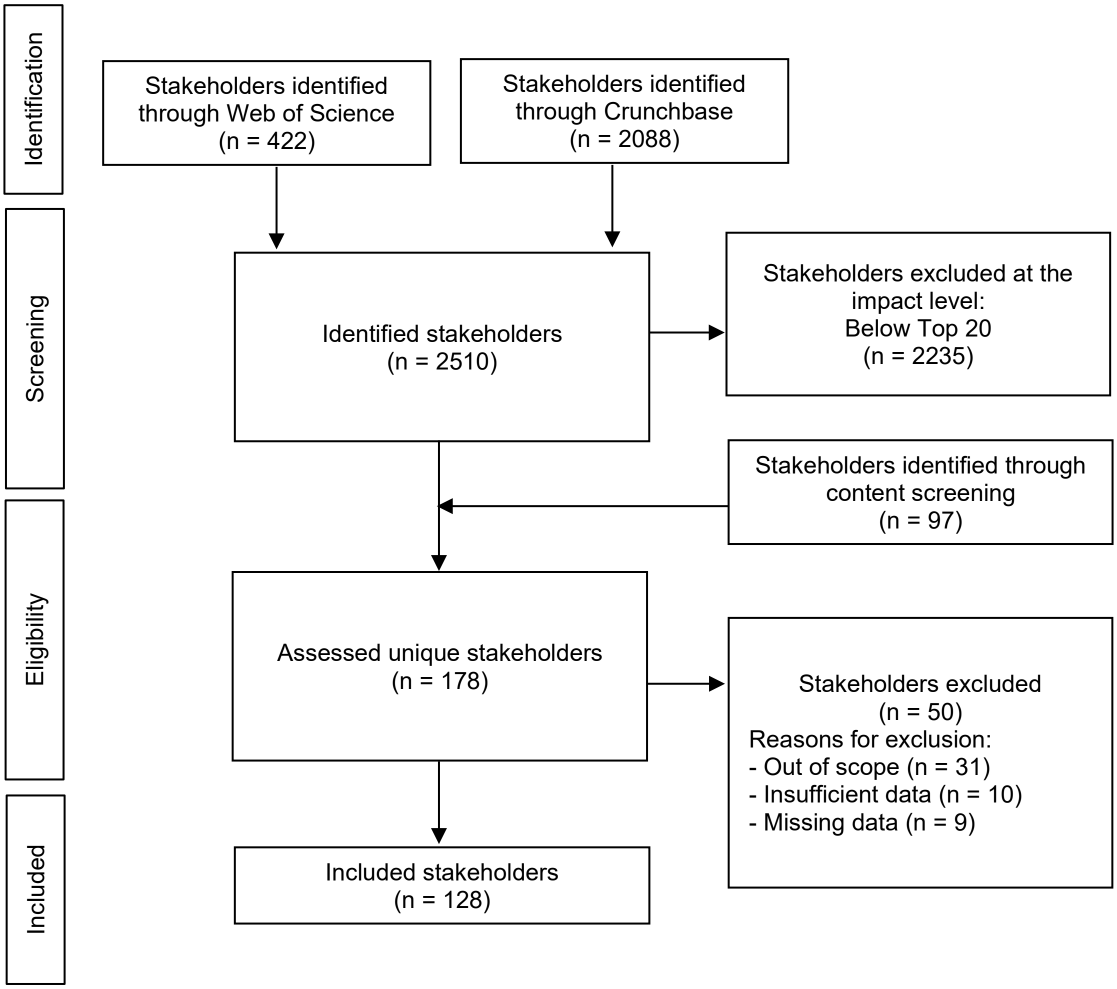


*Modeling and analyses*

*RQ1: Value proposition identification*. We used a natural language processing technique, semantic topic modeling, to identify the main themes of stakeholders' value proposition statements, addressing RQ1. Specifically, we employed Latent Dirichlet Allocation (LDA), a widely used computational technique that automatically identifies latent topics within a text corpus (Jelodar *et al.*, 2019). LDA allows for the discovery of recurring themes by analyzing the distribution of words across documents. We implemented LDA using the TopicModels and TidyText packages in R Studio (Silge and Robinson, 2017).

Following LDA standards in the field, we implemented several pre-processing steps (Silge and Robinson, 2017). First, stop words, such as "the," "and," and "in," were removed from the text corpus to focus on more meaningful content. Additionally, punctuation and special characters were stripped to ensure consistency in text processing. Next, the text was tokenized, breaking it down into individual words. Words were then stemmed or lemmatized to reduce them to their base form, thereby standardizing variations of the same word. Finally, the preprocessed text was converted into a document-term matrix, with each cell indicating the frequency of a word in a document. This matrix served as input for the LDA algorithm to identify latent topics within the text corpus.

We filtered the top words from this text corpus that appeared more than once. In optimizing the number of resulting topic allocations, we utilized coherence scores to determine the most suitable number of topics from a possible range of 2-4. Coherence scores evaluate the interpretability and consistency of topics generated by LDA by calculating the co-occurrence of words within the same context window across the corpus. Higher coherence scores indicate that the words within each topic are more semantically similar and coherent. Thus, the model with the optimal number of topics was selected based on the output with the highest coherence score, ensuring that the generated topics were meaningful and coherent. This approach allowed us to fine-tune the LDA model best to capture the underlying themes in the stakeholder value propositions. For the resulting topic categorizations, see tables S2-S7.

*RQ2: Value proposition alignment.* Next, we took a network analysis approach to assess the level of agreement between the value propositions or proposed initiatives among key stakeholders (RQ2). Network analysis helps assess the level of agreement between stakeholders' value propositions as it allows for representing complex relationships and provides quantitative measures to evaluate alignment. By quantifying alignment using metrics like cosine similarity, we can identify central stakeholders and understand the dynamics of collaboration in the healthy longevity landscape (Wasserman and Faust, 1994).

We first constructed an undirected network, where stakeholders are represented as nodes, and the strength of value proposition alignment serves as weighted edges. Each stakeholder is connected to others based on the degree of alignment observed between their value propositions. To quantify the level of alignment among stakeholders' value propositions, we utilized cosine similarity, a popular semantic similarity metric in text analyses (Silge and Robinson, 2017) similarity measures the cosine of the angle between two vectors, representing the similarity between the value propositions of different stakeholders. Higher cosine similarity values indicate greater alignment, suggesting the stakeholders' value propositions share similar themes. The cosine similarity between two vectors $A$ and B is computed as:

$$\text{cosine\_similarity}(\mathbf{A}, \mathbf{B}) = \frac{\sum_i A_i \cdot B_i}{\sqrt{\sum_i A_i^2} \cdot \sqrt{\sum_i B_i^2}}$$

Next, we calculated the normalized node strength for each stakeholder to assess the alignment of their value propositions with those of other stakeholders in the network. By considering the weighted sum of cosine similarities per stakeholder, we gain insights into the extent to which each stakeholder's value propositions align with the collective themes or content present in the network. Normalized node strength for a stakeholder node $i$ is calculated as:

$$\text{Normalized Node Strength}(i) = \frac{\sum_j \text{cosine\_similarity}(i,j)}{\sum_j \text{Total Weighted Edges}(i,j)}$$

Where $j$ represents other stakeholder nodes connected to node $i$. Total Weighted Edges $(i,j)$ denotes the total cosine similarity between node $i$ and node $j$. The normalized node strength measures the relative alignment of each stakeholder value proposition within the network based on the alignment of their value propositions with those of other stakeholders.

A normalized node strength close to 1 indicates that a stakeholder's value propositions are highly aligned with those of other stakeholders in the network, suggesting strong coherence. On the other hand, a normalized node strength closer to 0 suggests lower alignment with other stakeholders.

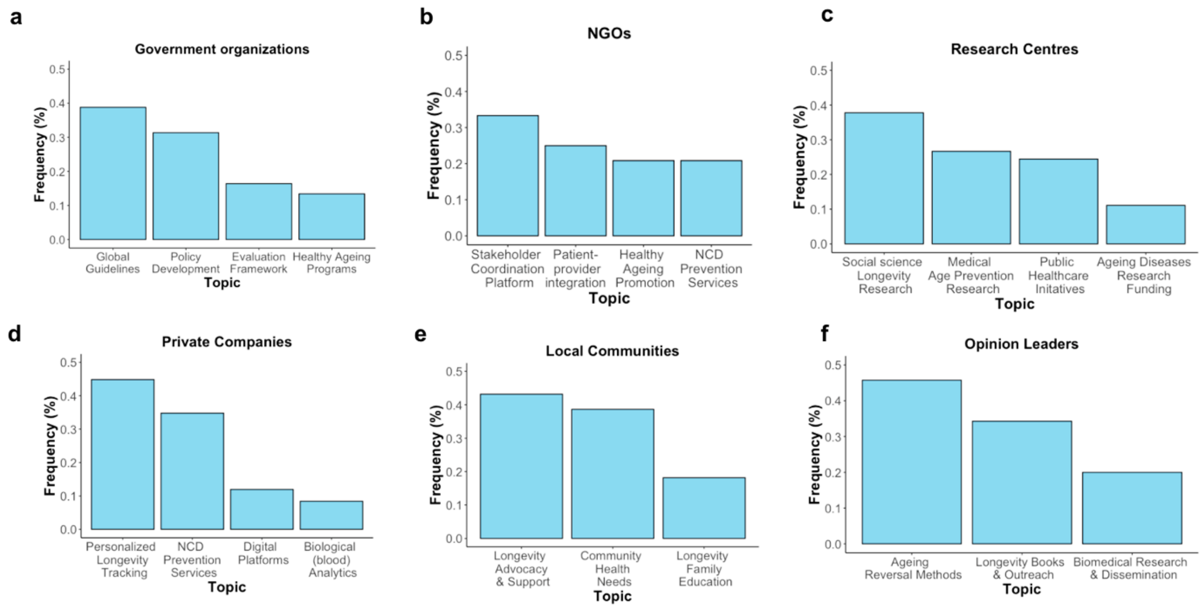


Figure S2. Key topics identified in stakeholders' healthy longevity related value proposition statements (*N=128*) using semantic topic modeling.

#### SUPPLEMENTAL REFERENCES


Bautmans, I. *et al.* (2022) 'WHO working definition of vitality capacity for healthy longevity monitoring', *The Lancet Healthy Longevity*, 3(11), pp. e789–e796. Available at: https://doi.org/10.1016/S2666-7568(22)00200-8.

Cozzolino, A. and Geiger, S. (2024) 'Ecosystem disruption and regulatory positioning: Entry strategies of digital health startup orchestrators and complementors', *Research Policy*, 53(2), p. 104913. Available at: https://doi.org/10.1016/j.respol.2023.104913.

Crunchbase (2024) 'Crunchbase Inc.' Available at: https://www.crunchbase.com/home (Accessed: 7 May 2024).

Ebadi, A. and Schiffauerova, A. (2016) 'How to boost scientific production? A statistical analysis of research funding and other influencing factors', *Scientometrics*, 106(3), pp. 1093–1116. Available at: https://doi.org/10.1007/s11192-015-1825-x.

Fernandez-Moure, J.S. (2016) 'Lost in Translation: The Gap in Scientific Advancements and Clinical Application', *Frontiers in Bioengineering and Biotechnology*, 4. Available at: https://doi.org/10.3389/fbioe.2016.00043.

Fried, L.P., Wong, J.E.-L. and Dzau, V. (2022) 'A global roadmap to seize the opportunities of healthy longevity', *Nature Aging*, pp. 1–4. Available at: https://doi.org/10.1038/s43587-022-00332-7.

Gassmann, O., Frankenberger, K. and Choudury, M. (2020) *The Business Model Navigator: 55+ models that will revolutionise your business*. Second Edition. New York: Pearson.

IHME (2023) 'Financing Global Health'. University of Washington. Available at: https://vizhub.healthdata.org/fgh/.

Jelodar, H. *et al.* (2019) 'Latent Dirichlet allocation (LDA) and topic modeling: models, applications, a survey', *Multimedia Tools and Applications*, 78(11), pp. 15169–15211. Available at: https://doi.org/10.1007/s11042-018-6894-4.

Lingens, B., Seeholzer, V. and Gassmann, O. (2022) 'The architecture of innovation: how firms configure different types of complementarities in emerging ecosystems', *Industry and Innovation*, 29(9), pp. 1108–1139. Available at: https://doi.org/10.1080/13662716.2022.2123307.

National Academy of Medicine (2022) *Global Roadmap for Healthy Longevity*. Washington, D.C.: National Academies Press, p. 26144. Available at: https://doi.org/10.17226/26144.

Payne, A. and Frow, P. (2014) 'Developing superior value propositions: a strategic marketing imperative', *Journal of Service Management*. Edited by P. Bo Edvardsson And Professor Philipp Klaus, 25(2), pp. 213–227. Available at: https://doi.org/10.1108/JOSM-01-2014-0036.

PitchBook (2024) 'Venture Capital, Private Equity and M&A Database'. Available at: https://pitchbook.com/ (Accessed: 7 May 2024).

Silge, J. and Robinson, D. (2017) *Text mining with R: a tidy approach*. First edition. Beijing Boston Farnham Sebastopol Tokyo: O'Reilly.

Steinhöfel, E., Kohl, H. and Orth, R. (2016) *Business Model Innovation: A Comparative Analysis*.

Valdez, D., Pickett, A.C. and Goodson, P. (2018) 'Topic Modeling: Latent Semantic Analysis for the Social Sciences', *Social Science Quarterly*, 99(5), pp. 1665–1679. Available at: https://doi.org/10.1111/ssqu.12528.

Wasserman, S. and Faust, K. (1994) *Social Network Analysis: Methods and Applications*. Cambridge: Cambridge University Press (Structural Analysis in the Social Sciences). Available at: https://doi.org/10.1017/CBO9780511815478.

WHO (2020) *Decade of healthy ageing: baseline report*. Geneva: World Health Organization. Available at: https://apps.who.int/iris/handle/10665/338677 (Accessed: 12 September 2022).

WHO (2023) *Non communicable diseases Key Facts*. Available at: https://www.who.int/news-room/fact-sheets/detail/noncommunicable-diseases (Accessed: 12 March 2024).
